# Robust immunogenicity of a third BNT162b2 vaccination against SARS-CoV-2 Omicron variant in a naïve New Zealand cohort

**DOI:** 10.1101/2023.03.30.23287981

**Authors:** Brittany Lavender, Caitlin Hooker, Chris Frampton, Michael Williams, Simon Carson, Aimee Paterson, Reuben McGregor, Nicole J. Moreland, Katie Gell, Frances H. Priddy, Kjesten Wiig, Graham Le Gros, James E. Ussher, Maia Brewerton

**Author notes:** Corresponding Author: Dr Maia Brewerton or Brittany Lavender, Address: Malaghan Institute of Medical Research, PO Box 7060, Wellington 6242, New Zealand.

## Abstract

The ability of a third dose of the Pfizer-BioNTech BNT162b2 SARS-CoV-2 vaccine to stimulate immune responses against subvariants, including Omicron BA.1, has not been assessed in New Zealand populations. Unlike many overseas populations, New Zealanders were largely infection naïve at the time they were boosted. This adult cohort of 298 participants, oversampled for at-risk populations, was composed of 29% Māori and 28% Pacific peoples, with 40% of the population aged 55+. A significant proportion of the cohort was obese and presented with at least one comorbidity. Sera were collected 28 days and 6 months post second vaccination and 28 days post third vaccination. SARS-CoV-2 anti-S IgG titres and neutralising capacity using surrogate viral neutralisation assays against variants of concern, including Omicron BA.1, were investigated. The incidence of SARS-CoV-2 infection, within our cohort, prior to third vaccination was very low (<6%). This study found a third vaccine significantly increased the mean SARS-CoV-2 anti-S IgG titres, for every demographic subgroup, by a minimum of 1.5-fold compared to titres after two doses. Diabetic participants experienced a greater increase (∼4-fold) in antibody titres after their third vaccination, compared to non-diabetics (increase of ∼2-fold). This corrected for the deficiency in antibody titres within diabetic participants which was observed following two doses. A third dose also induced a neutralising response against Omicron variant BA.1, which was absent after two doses. This neutralising response improved regardless of age, BMI, ethnicity, or diabetes status. Participants aged ≥75 years consistently had the lowest SARS-CoV-2 anti-S IgG titres at each timepoint, however experienced the greatest improvement after three doses compared to younger participants. This study shows that in the absence of prior SARS-CoV-2 infection, a third Pfizer-BioNTech BNT162b2 vaccine enhances immunogenicity, including against Omicron BA.1, in a cohort representative of at-risk groups in the adult New Zealand population.

## Introduction

It has been over three years since Severe Acute Respiratory Syndrome Coronavirus 2 (SARS-CoV-2) first emerged in Wuhan, China, with the World Health Organization (WHO) declaring this novel coronavirus a public health emergency in January 2020 [1]. As SARS-CoV-2 spread rapidly around the world causing coronavirus disease (COVID-19), Aotearoa New Zealand introduced stringent public health measures in pursuit of a SARS-CoV-2 elimination strategy, in March 2020 [2]. This strategy prevented the widespread community transmission of SARS-CoV-2 in New Zealand before effective COVID-19 vaccines were available to the public early in 2021. Therefore, prior to the introduction of Omicron and the termination of the elimination strategy in early 2022, incident SARS-CoV-2 infections were rare in the wider New Zealand population. This provided a unique opportunity to study the immunogenicity of a third vaccine in the absence of wide-spread infection.

Until January 2022, when the highly transmissible Omicron variant entered the community [3], New Zealand experienced very low rates of SARS-CoV-2 infection (<0.3% of the total population), despite a controlled outbreak of the Delta variant in the Auckland region [4]. The Omicron outbreak began with the BA.1 variant which was quickly overtaken by BA.2 [5, 6]. At the time of writing, the CH.1.1 and XBB variants are the most prevalent Variant of Concerns (VoC) within New Zealand [7]. After the Omicron variant became established in New Zealand, the epidemiology of COVID-19 mortality risk mirrored what had been seen overseas, with increasing age being the strongest risk factor for death from COVID-19 in New Zealand [8, 9]. Inequities in COVID-19 mortality risk were seen as Māori and Pacific peoples experienced more than double the mortality risk compared to other ethnicities. If these individuals had at least one comorbidity, their risk increased 6-fold compared to those without comorbidities [9].

New Zealand’s nationwide COVID-19 immunisation programme began in February 2021 [3], with a stepwise vaccination roll out according to eligibility criteria. The majority of New Zealander’s became eligible for vaccination by July 2021 [4]. Pfizer-BioNTech BNT162b2 is the preferred COVID-19 vaccine in New Zealand with it comprising 99.9% (11,900,035 out of 11,916,152) of administered vaccine doses. NovaVax NVX-CoV2373 and Oxford/AstraZeneca ChAdOx1 nCoV-19 doses comprise the small remainder [5].

New Zealand achieved very high uptake of the COVID-19 vaccine with 89.3% of those aged 12 years and over having received two doses [5]. In line with the results of international data, in November 2021 a third dose was recommended by the New Zealand Government. This was initially recommended to be administered 6 months after the primary two-dose schedule but, due to increasing community transmission in February 2022, was brought forward to 4 months and then 3 months shortly after [6-8]. The uptake of the third dose has been significantly lower with 73.2% of New Zealanders aged 18 years and over receiving at least three doses [9].

In this study, we present further immunogenicity findings following BNT162b2 vaccination from the ongoing cohort study, Ka Mātau, Ka Ora, conducted in New Zealand. This cohort includes some of those at higher risk of severe disease or death from COVID-19 including older people, Māori, Pacific peoples, and those with co-morbidities. Prior results from this cohort showed an absence of neutralising capacity of antibodies against Omicron (BA.1) 28 days after two BNT162b2 vaccine doses [10]. Additionally, participants with diabetes showed reduced immunogenicity against SARS-CoV-2 following two doses compared to non-diabetics [10]. Here we evaluate waning of antibody responses 6 months after two doses of BNT162b2 and the impact of a third dose on antibody responses in a largely SARS-CoV-2 infection naïve cohort while comparing across age groups, ethnicities, and co-morbidities.

## Methods

### Study Design and Population

In this ongoing cohort study, Ka Mātau, Ka Ora, a total of 298 adult participants were enrolled at study commencement in June 2021. There is an enrichment of at-risk populations in New Zealand in the cohort, including Māori, Pacific peoples, adults over 65 years of age, and those with comorbidities associated with a greater risk of severe COVID-19 disease. Comorbidities recorded include cardiac, pulmonary, endocrine and haematologic conditions. Participants with primary or secondary immunodeficiencies were excluded, and key inclusion and exclusion criteria are as published [10]. Participant recruitment occurred outside of New Zealand’s most populated area (the Auckland region) at two Pacific Clinical Research Network locations in Rotorua (North Island) and Christchurch (South Island). Participants were administered two primary doses, and a third dose of the Pfizer-BioNTech BNT162b2 vaccine, between June 2021 and April 2022, in accordance with New Zealand’s national COVID-19 immunisation programme. Blood serum samples were collected 28 days after the second dose, 6 months after the second dose (and prior to the third dose therefore this collection was brought forward in participants who received their third dose between 3 and 6 months) and 28 days post third vaccine dose. Vaccination dates were self-reported by participants and confirmed on vaccination cards where available.

### Ethical Approval

The New Zealand Health and Disability Ethics Committee granted approval for the original design of the Ka Mātau, Ka Ora Study and its analyses in June 2021 (21/NTB/117). Approval of amendments to the study design incorporating third vaccine doses were granted in December 2021.

### Demographic Variables

Key variables analysed in this cohort were age, gender, ethnicity, body mass index (BMI), and comorbidities, including diabetes status. Age was defined at time of enrolment based on year of birth. Participant age was not altered as part of the longitudinal study, which continued over an approximate 12-month period. Prioritised ethnicity was self-reported by participants using the categories provided as part of the New Zealand Ethnicity Protocols [11]. Participant BMIs were categorised in accordance with the New Zealand Ministry of Health guidelines: 18.5 kg/m^2^ - 24.9 kg/m^2^ (healthy weight), 25.0 kg/m^2^ - 29.9 kg/m^2^ (overweight) and ≥ 30.0 kg/m^2^ (obese) [12].

### Laboratory Analysis

#### Anti-SARS-CoV-2- nucleocapsid specific antibodies (including IgG)

Serum samples were analysed at Southern Community Laboratories in Dunedin (New Zealand). The electrochemiluminescence Elecsys® Anti-SARS-CoV-2 immunoassay (Roche Ltd) was used, in accordance with manufacturer instructions, to detect antibodies that recognise SARS-CoV-2 nucleocapsid protein in serum, which indicates prior infection [13]. Participants were tested at baseline (pre vaccination), prior to third dose and 28 days post third vaccine dose. The manufacturer’s cut-off for a reactive sample (COI > 1) was used.

#### SARS-CoV-2 anti-S IgG antibodies

Binding antibodies specific to the Ancestral SARS-CoV-2 spike protein were quantified using the Abbott SARS-CoV-2 IgG II Quant immunoassay (Abbott Laboratories, Abbott Park, USA). This was conducted at Southern Community Laboratories, according to manufacturer’s instructions as previously described [13]. The manufacturer’s cut off for a positive sample remains as 50 AU/mL. Quantitative titres were converted to WHO International Units using a conversion factor supplied by the manufacturer.

#### Neutralising SARS-CoV-2 antibodies

Neutralising antibodies against Ancestral, Beta, Delta, and Omicron BA.1 variants of SARS-CoV-2 were quantified using the cPass SARS-CoV-2 Surrogate Virus Neutralisation Test (sVNT) (Genscript, Singapore), in accordance with the manufacturer’s guidelines at the University of Auckland (New Zealand). This assay is validated against live viral neutralisation assays, and measures reduction in binding of recombinant spike and ACE2 proteins as a surrogate for neutralisation of viral infection [13, 14]. Results were reported as percent (%) inhibition for all VoC. A value below 20% indicates a negative result.

#### Statistical Methods

Anti-S IgG are reported as Geometric Mean Titres (GMT). Geometric Mean Fold Rise (GMFR) describes the change in GMT of anti-S IgG between two different time points. Values were adjusted for demographic variables, BMI, age, ethnicity, and diabetes. Change (Δ) in % inhibition between 6 months post second dose and 28 days post third dose was calculated for the sVNT assay results.

All participants with datapoints were included in the analysis for each timepoint and each variable measured, with no missing data imputation for sporadic missing assay values. Sample sizes and demographic features therefore differed slightly depending on the timepoint and assay values analysed, as indicated within the descriptive tables. The immunogenicity and neutralisation values were transformed prior to analysis to normalise distributions and analysed using general linear models, including age, BMI, ethnicity, and diabetes as covariates. Tabled results represent model-derived, back-transformed means and 95% confidence intervals. The percentage of participants with neutralisation against SARS-CoV-2 variants were analysed using a multivariable logistic regression model, with age, BMI, ethnicity, and diabetes as covariates. All analyses were undertaken using SPSS v28, and a two-tailed p-value <0.05 was taken to indicate statistical significance.

#### Figures and Tables

Graphical visualisations were created using Prism software (version 9.4.1). Antigenic cartography maps were created using the Racmacs function (version 4.2.1) in R Studio, with inhibition data transformed to mimic titre data.

## Results

### Study Participants

From 331 participants who were screened, the study enrolled 298 participants. A small number of participants were excluded after enrollment (n = 2) or lost to follow up (n = 8) at different timepoints (Figure 1). Twenty-eight days after second vaccination, 288 participants remained enrolled with 287 samples analysed [13]. At 6 months post second dose (prior to third dose), 273 participants remained in the study with immunogenicity analysis performed on 239 (anti-S IgG analysis and anti-nucleocapsid testing) and 236 (surrogate viral neutralisation testing) participants, respectively. At 28 days post third dose, 236 of the remaining 257 participants had their samples analysed on all three assays (Figure 1). The average duration of time from enrolment to serum collection at the 28-day post third dose timepoint was 238 days (approximately 8 months); this ranged from 174 days to 354 days.

**Figure 1.**
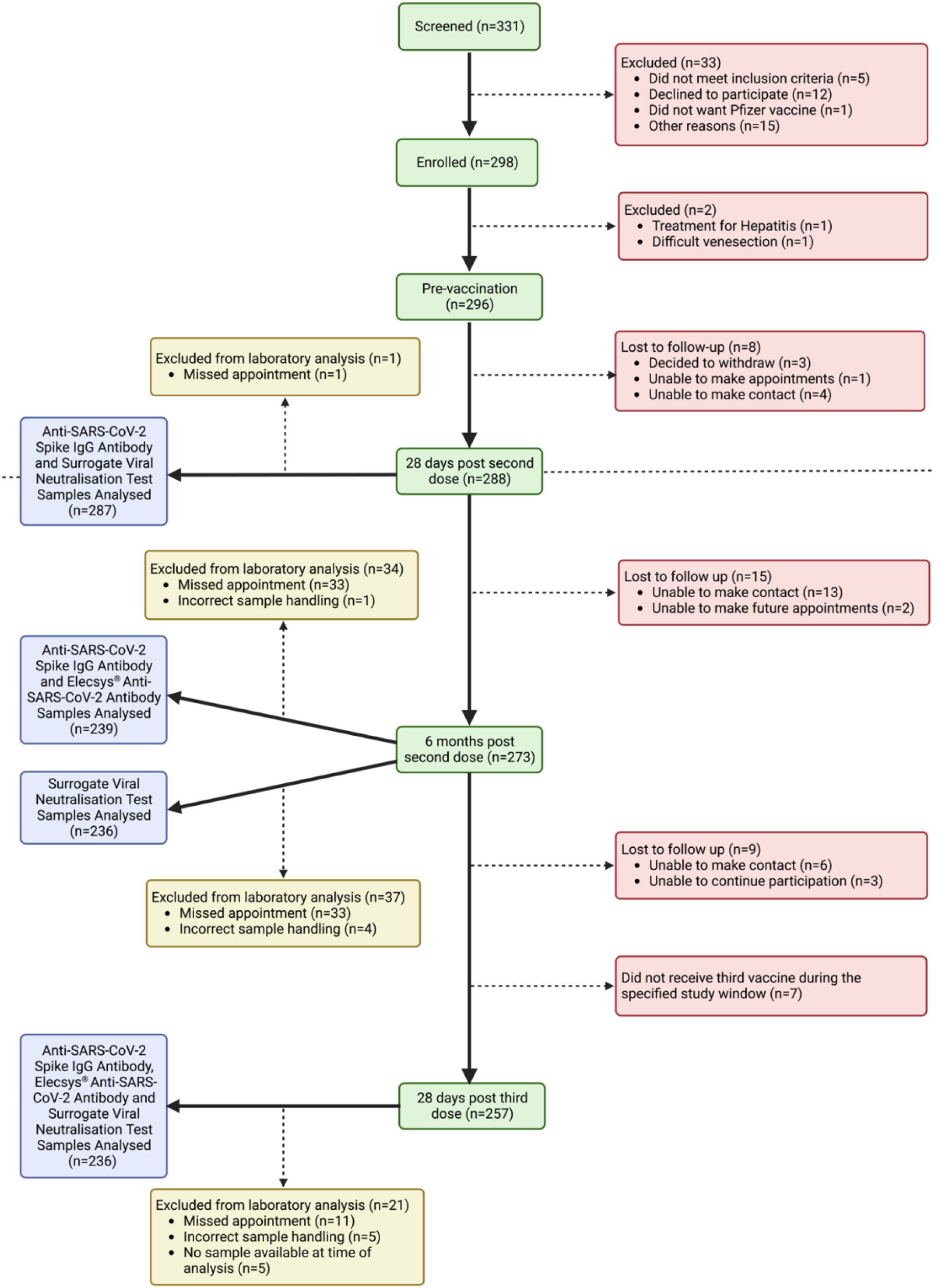
Participant flow-through of the Ka Mātau, Ka Ora study to date. Created with BioRender.com.

### Demographics

The study population includes several at risk New Zealand populations, with demographics summarised in Table 1. Māori and Pacific populations comprise 29.9% and 28.1% of the study population compared to approximately 17% and 8% of the total New Zealand population, respectively [15]. Half of the Pacific study population are Samoan, with the second most prevalent Pacific ethnicity being Tongan (13.6%). New Zealand European represent the largest proportion of non-Māori, non-Pacific participants making up 34.7% of the total study population. Māori and Pacific participants have a lower median age (50 years and 42 years, respectively) compared to 70 years for New Zealand European. Almost 45% of the study population are aged over 54 years, with 16.3% being 75 years or older. The BMI of participants ranges from 18.8 kg/m^2^ to 61.0 kg/m^2^; with 16% of participants having a healthy body weight and over half of the study population being classified as obese (55.9%). 10% of the study population are diabetic, with type 2 diabetes being the most prevalent (Table 1).

**Table 1.** Demographics of all participants included in analysis at any timepoint.

|  | % (n) | Median age (IQR) |
| --- | --- | --- |
| *Total Participants* | 288 | 52 (35 – 66) |
| *Gender* |  |  |
| Female | 54.2 (156) | 50 (34 – 65) |
| Male | 45.5 (131) | 54 (39 – 66) |
| Non-Binary | 0.3 (1) | 31 |
| *Age (years)* |  |  |
| 16-34 | 25.0 (72) | - |
| 35-54 | 30.6 (88) | - |
| 55-64 | 19.1 (55) | - |
| 65-74 | 9.0 (26) | - |
| 75+ | 16.3 (47) | - |
| *Ethnicity* |  |  |
| Non-Māori and Non-Pacific Peoples | 42.0 (121) | 60 (49 – 76) |
| NZ European | 82.6 (100) | 70 (49 – 77) |
| Other | 17.4 (21) | 54 (46 – 59) |
| Māori | 29.9 (86) | 50 (35 – 59) |
| Pacific Peoples | 28.1 (81) | 42 (26 – 52) |
| Samoan | 56.8 (46) | 43 (26 – 56) |
| Tongan | 13.6 (11) | 32 (21 – 45) |
| Tokelauan | 11.1 (9) | 49 (37 – 55) |
| Cook Island Māori | 8.6 (7) | 48 (32 – 52) |
| Fijian | 6.2 (5) | 36 (28 – 47) |
| Other | 3.7 (3) | 38 (38 – 41) |
| *BMI* |  |  |
| Healthy | 16.0 (46) | 49 (23 – 67) |
| Overweight | 28.1 (81) | 54 (38 – 63) |
| Obese | 55.9 (161) | 51 (38 – 63) |
| *Diabetes Status* |  |  |
| Present | 9.7 (28) | 58 (50 – 72) |
| Absent | 90.3 (260) | 50 (33 – 63) |

Other non-Māori and non-Pacific peoples includes European not further defined (8), British (7), South African (3), Dutch (2), Middle Eastern/Latin American/African (1).
Other Pacific peoples includes Kiribati (1), Tuvalu (1), Cook Island Māori/Samoan (1).
Healthy BMI 18.5 kg/m<sup>2</sup> - 24.9 kg/m<sup>2</sup>; Overweight BMI 25.0 kg/m<sup>2</sup> - 29.9 kg/m<sup>2</sup>; Obese BMI ≥ 30.0 kg/m<sup>2</sup>.
Diabetes type 1 (2), type 2 (26).

### Third Dose Vaccination and Laboratory Analysis Windows

Participants received their third vaccination between 3.0 and 7.8 months after their second dose, with a median of 5.7 months (Supplementary Figure 1). The collection of samples at the 28 days post third dose timepoint ranged from 4 to 160 days post third dose. The sample collection had a preferred window of 21-56 days post third dose, with 85% percent of participants being within this window, with a median of 32 days. All samples (n = 4) collected <21 days post third dose were excluded from analysis (Supplementary Figure 2). Variations in vaccination window or sample collection window (after 21 days) did not have statistically significant effects on immunogenicity of binding antibody titres or neutralisation responses in this cohort (Supplementary Table 2). However, this study was not powered to detect these temporal differences.

### SARS-CoV-2 prior infection: nucleocapsid seropositivity

At the time of sample analysis, there were very low rates of prior SARS-COV-2 infection in this cohort. Three (1.3%) participants tested positive for anti-nucleocapsid antibodies prior to their third dose, which increased to 11/236 (4.6%) 28 days post third dose. Statistical analysis showed the inclusion of these participants did not significantly impact primary outputs of analysis (data not shown) and were included in all subsequent analyses.

### SARS-CoV-2 Ancestral Anti-S IgG antibodies

In the total study population, anti-S IgG GMT waned approximately 10-fold from 966 IU/mL 28 days post second dose to 101 IU/mL 6 months post-second vaccine dose. This waning was statistically significant and seen across all ethnicities, BMI, and age subgroups (p <0.001, Figure 2). Comparing GMT 28 days post second dose to 28 days post third dose, there was an increase from 966 IU/mL to 1892 IU/mL, equating to an almost 2-fold rise in anti-S IgG antibodies following a third dose (p <0.001, Figure 2).

**Figure 2.**
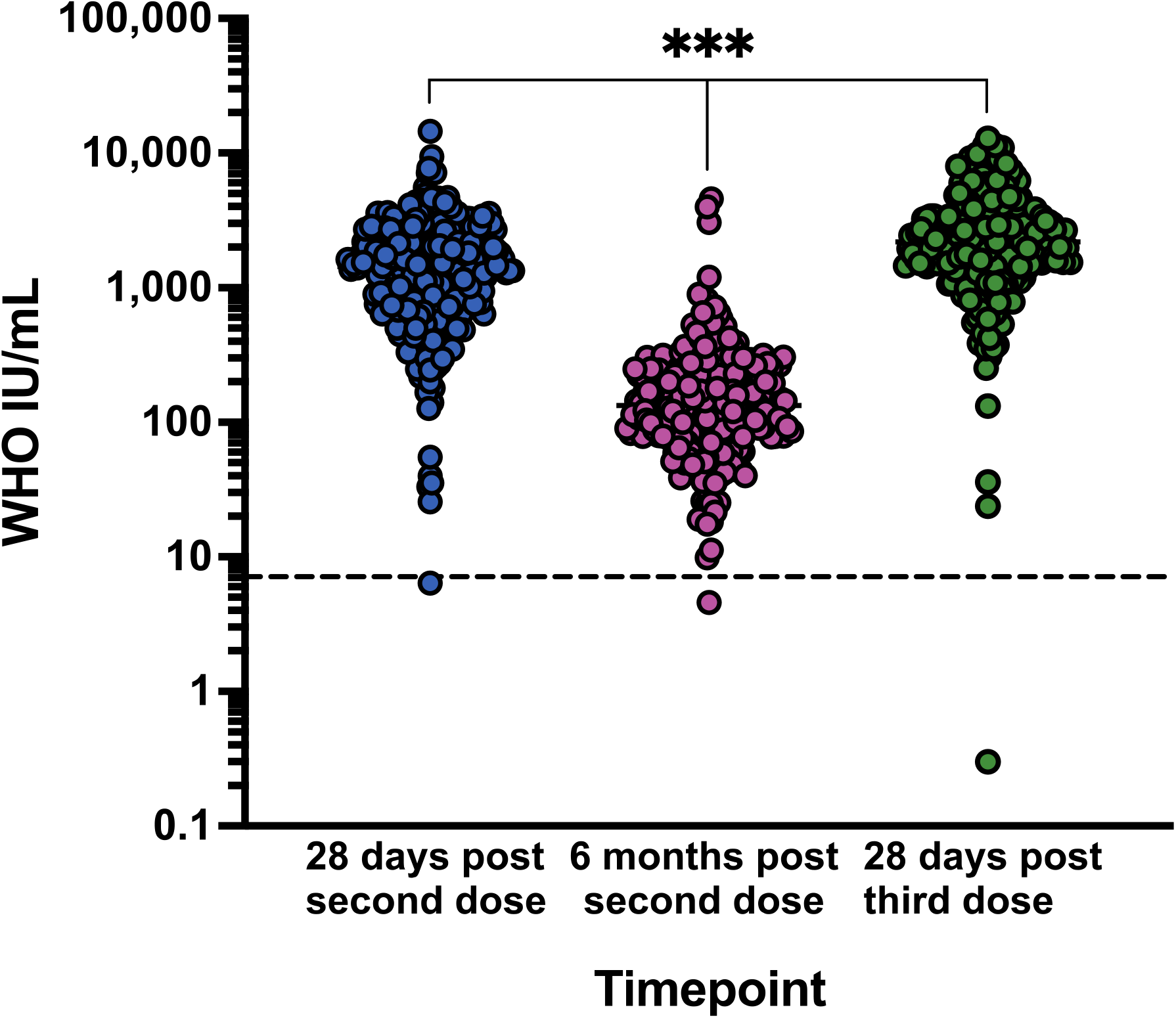
Unadjusted SARS-CoV-2 anti-S IgG antibody titres, for all study participants after two and three doses of BNT162b2 vaccine. Cut off for positive result is 7.1 IU/mL (horizontal dotted line). *** p < 0.001 comparing each group to each other

All age groups had significant improvements in antibody titre from the third vaccine dose, compared to 28 days after second dose (p < 0.001 for all, Figure 3A). After a third vaccination, 16-34-year-olds GMT increased almost 2-fold (1520.2 IU/mL to 2939.7 IU/mL), 35-54-year-olds increased 1.5-fold (1188.1 IU/mL to 1848.4 IU/mL), 55-64-year-olds increased 1.7-fold (1005 IU/mL to 1717.5 IU/mL), 65-74-year-olds increased 2.8-fold (824.3 IU/mL to 2318.1 IU/mL) and 75+ year-olds increased 2.8-fold (421.3 IU/mL to 1175 IU/mL, Figure 3A). When comparing the GMFR, (the level of SARS-CoV-2 anti-S IgG 6 months post second dose relative to 28 days post third dose), participants aged 75 years and older showed the greatest improvement (highest GMFR) after three doses. This difference was statistically significant compared to three other ages groups, 16-34-year-olds (p = 0.022), 35-54-year-olds (p = 0.002) and 55-64-year-olds (p = 0.016), but not compared to 65-74-year-olds (Table 2). All age data was adjusted for BMI, ethnicity, and diabetes status.

**Figure 3.**
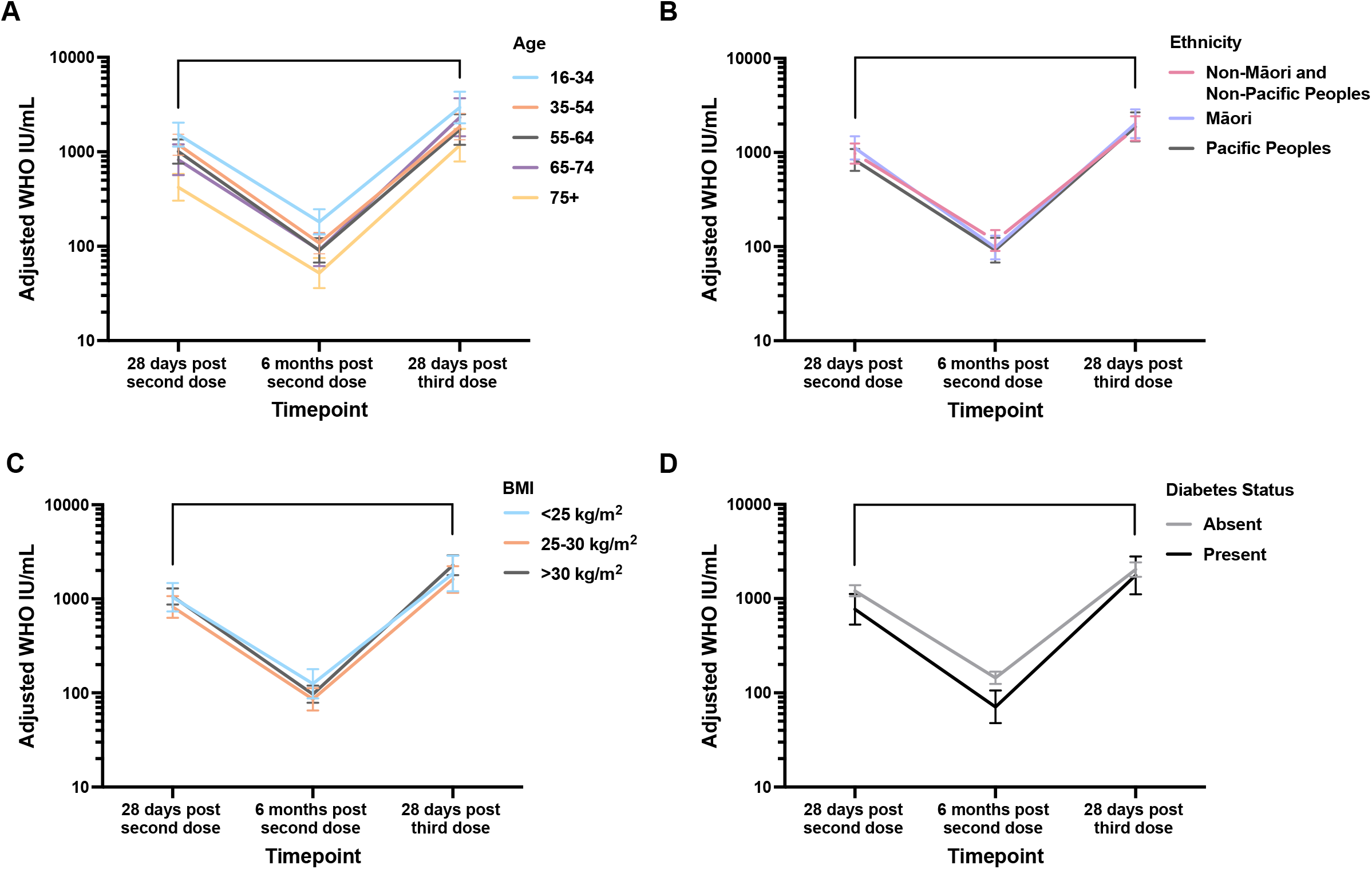
SARS-CoV-2 anti-S IgG geometric mean titres, for key demographic groups, after two and three doses of BNT162b2 vaccine, adjusted for age, BMI, ethnicity, and diabetes status (95% CI). **\*\*\*** p < 0.001 for all subgroups comparing timepoints indicated.

**Table 2.**
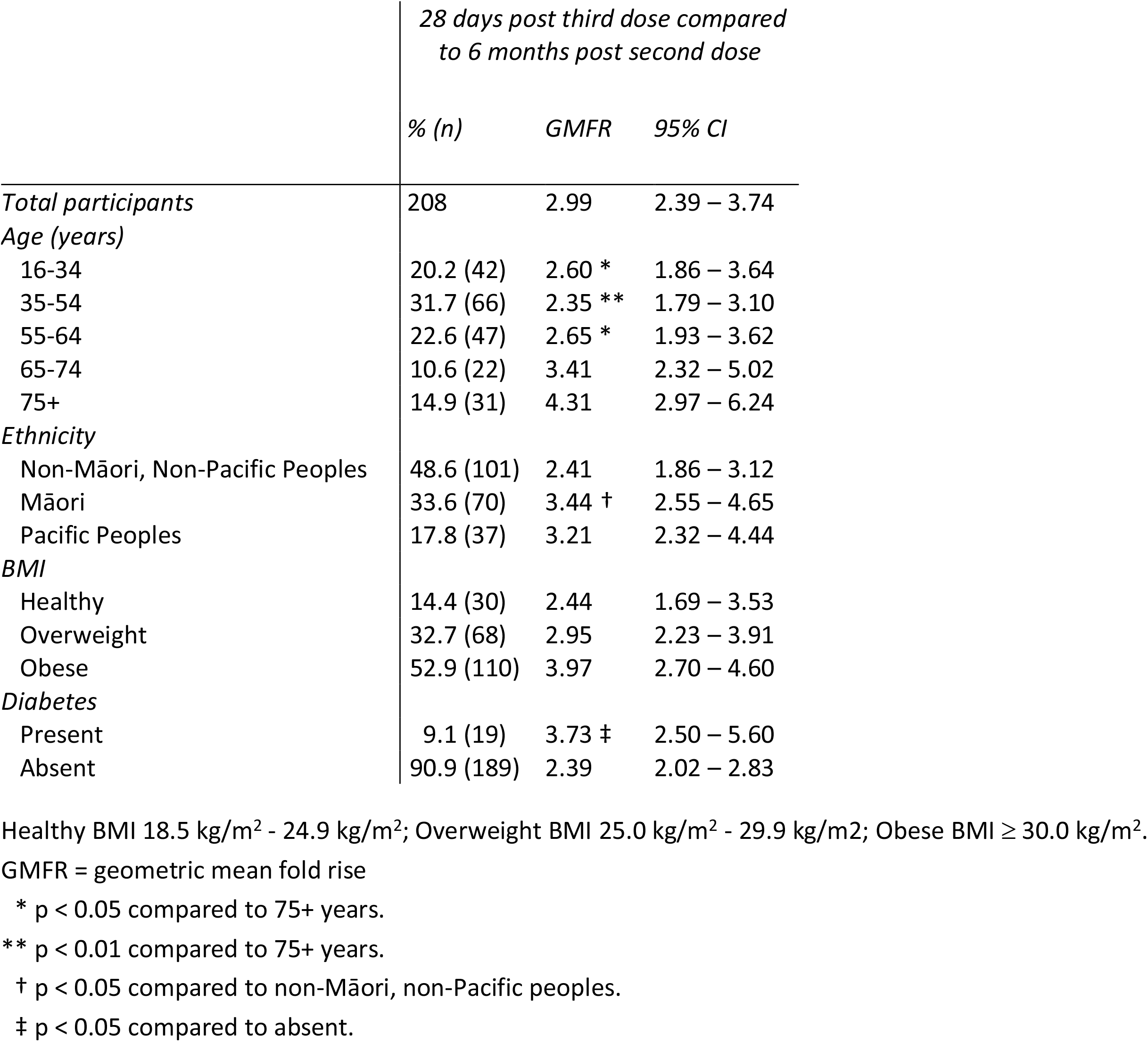
SARS-CoV-2 anti-S IgG antibody titre geometric mean fold rise after three doses of BNT162b2 vaccine, adjusted for age, ethnicity, BMI, and diabetes status.

A significant increase in GMT from 28 days post second dose to 28 days post third dose was seen across some ethnic groups in the study (p < 0.001, Figure 3B). Non-Māori and non-Pacific peoples and Māori both increased approximately 1.8-fold after three doses from 971.6 IU/mL to 1800.2 IU/mL and 1118 IU/mL to 2021.1 IU/mL, respectively. After three doses, Pacific peoples GMT increased 2.2-fold (832.3 IU/mL to 1871.8 IU/mL, Figure 3B). There was no statistically significant difference in GMT between ethnicities at 6 months post second dose (p = 0.308) or 28 days post third dose (p = 0.782). However, the GMFR (from 6 months post second dose relative to 28 days post third dose) was significantly higher in Māori (GMFR = 3.44) compared to non-Māori and non-Pacific participants (GMFR = 2.41, p = 0.0013, Table 2). All ethnicity data was adjusted for age, BMI, and diabetes status.

No significant difference was observed in the GMT between different BMI groups within the timepoints 6 months post second dose (p = 0.113), and 28 days post third dose (p = 0.083). However, for each BMI subgroup analysed a significant increase in GMT from 28 days post second dose to 28 days post third dose was seen (p < 0.001, Figure 3C). Participants with a BMI between 18.5 and 25.0 kg/m^2^ (healthy weight) had their GMT increase 1.8-fold from 1039.5 IU/mL 28 days post second dose to 1856.7 IU/mL 28 days post third dose. After three doses, GMT of overweight participants (BMI 25.0 kg/m^2^ – 30.0 kg/m^2^) increased approximately 2-fold (819.6 IU/mL to 1609.4 IU/mL). Similarly, participants with a BMI of >30.0 kg/m^2^ (obese) had GMT increase approximately 2.2-fold (1061.2 IU/mL to 2279 IU/mL) after three doses of BNT162b2 (Figure 3C). Consistently, the GMFR of the BMI subgroups comparing 6 months post second dose to 28 days post third dose did not show clear statistically significant differences (p = 0.05). All BMI data was adjusted for age, ethnicity, and diabetes status.

At 28 days and 6 months post second dose, diabetic participants showed significantly lower GMT compared to non-diabetics (p = 0.022 and p = 0.001, respectively). Interestingly, 28 days post third dose, there was no longer a statistical significance between GMT for participants with or without diabetes (p = 0.556). Diabetic participants had a 2.3-fold increase in GMT from 722.2 IU/mL 28 days post second dose to 1764.8 IU/mL 28 days post third dose (p < 0.001, Figure 3D). This fold increase is higher than that observed in non-diabetics, where there was a 1.7-fold increase in GMT (1210.9 IU/mL to 2035.8 IU/mL, p < 0.001, Figure 3D). The anti-S IgG GMFR in participants with diabetes, was significantly greater post third dose (∼4-fold change), compared to participants without (∼2-fold change, p = 0.042, Table 2). All diabetes data was adjusted for age, ethnicity, and BMI.

### Neutralising antibodies against SARS-CoV-2

Neutralisation of the Ancestral variant waned from a median of 95.4% 28 days post second dose to 85.7% 6 months post second dose (Figure 4A). Beta variant neutralisation waned from 86.4% to 55.5% and Delta variant from 96.0% to 73.8% 28 days post second dose and 6 months post vaccination, respectively (Figure 4B & C). The primary two dose schedule of the Pfizer-BioNTech BNT162b2 vaccine did not induce detectable levels of neutralisation against the SARS-CoV-2 Omicron BA.1 variant in most participants (Figure 4D). At 28 days post second dose, 12.9% of participants had detectable neutralisation against Omicron BA.1 which reduced to 4.2% 6 months post second dose (Supplementary Table 3). After the third dose, Omicron BA.1 neutralisation levels were detectable in 88.1% of participants (Figure 4D, Supplementary Table 3).

**Figure 4.**
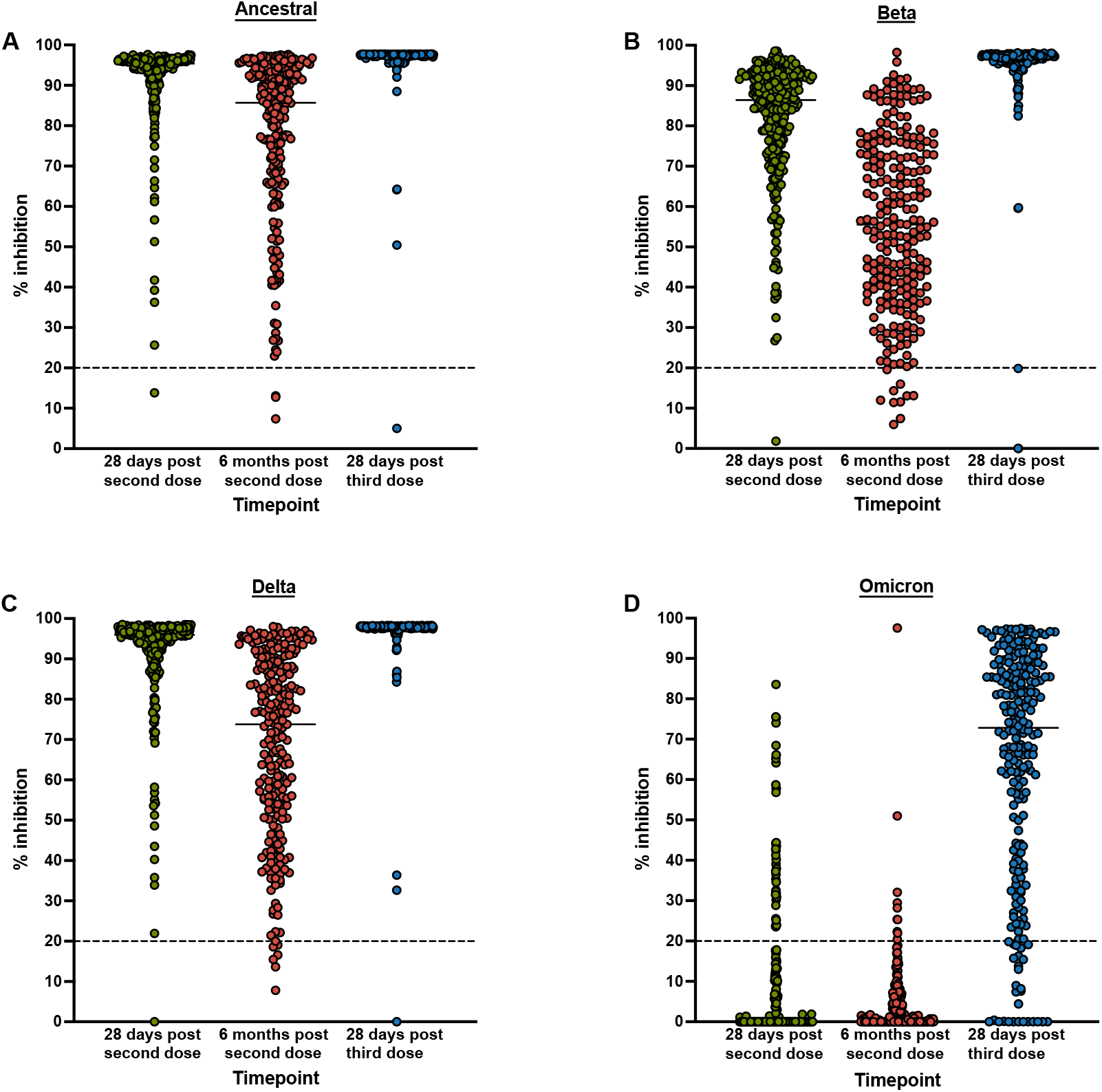
Unadjusted SARS-CoV-2 neutralising antibody responses to Ancestral, Beta, Delta and Omicron BA.1 variants after two and three doses of BNT162b2 vaccine. Cut-off for detectable result is 20% inhibition (horizontal dotted line).

Antigenic cartography illustrates the degree of antigenic difference between viral variants based on the antibody responses in sera (Figure 5). Following the third vaccination a convergence of antigens, particularly for Ancestral, Beta and Delta variants was observed. The antigenic difference between these three variants and Omicron BA.1 also reduced following the third dose, compared to after two doses. This is indicative of an increase in serum antibodies that cross-neutralise Omicron, following three doses of the vaccine compared with a primary (two dose) vaccination course.

**Figure 5.**
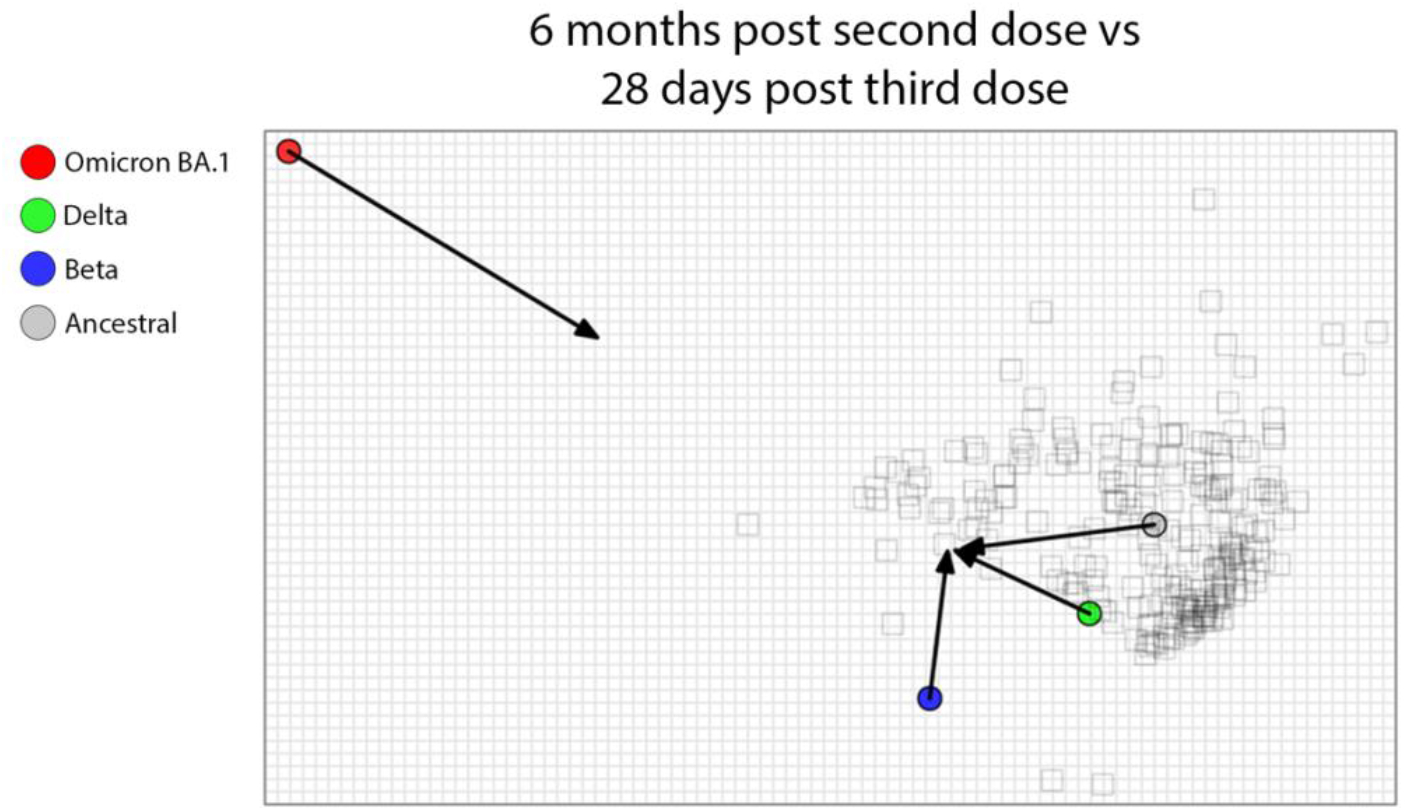
Antigenic cartography map displaying variants of concern (solid-coloured symbols) before and after three doses of BNT162b2 vaccine. Lined arrow distances between VoC (Ancestral, Beta, Delta, and Omicron BA.1) indicate shift in antigenic similarity using neutralising antibody (% inhibition). Individual sera are represented by hollow squares.

Given the predominance of circulating variants at the time this study was conducted, we examined the change in Omicron BA.1 neutralisation response in more detail. There was a significant increase in detectable Omicron BA.1 % inhibition from 6 months post second dose to 28 days post third dose, across the study population (Figure 6A-D, Table 3). Age was the only demographic variable for which the increase in Omicron BA.1 response differed. The mean neutralisation in 16-34-year-olds was significantly higher after three doses (82.7%) compared to 35-54-year-olds (72.3%, p = 0.016), 55-64-year-olds (71.1%, p = 0.016), 65-74-year-olds (70.5%, p = 0.043) and 75+ year-olds (63.0%, p = 0.001) (Table 3). Data was adjusted for ethnicity, BMI, and diabetes status. There was no statistically significant difference in mean Omicron BA.1 neutralisation response between different ethnicity groups, BMI ranges, or diabetes status after three vaccine doses (Table 3).

**Figure 6.**
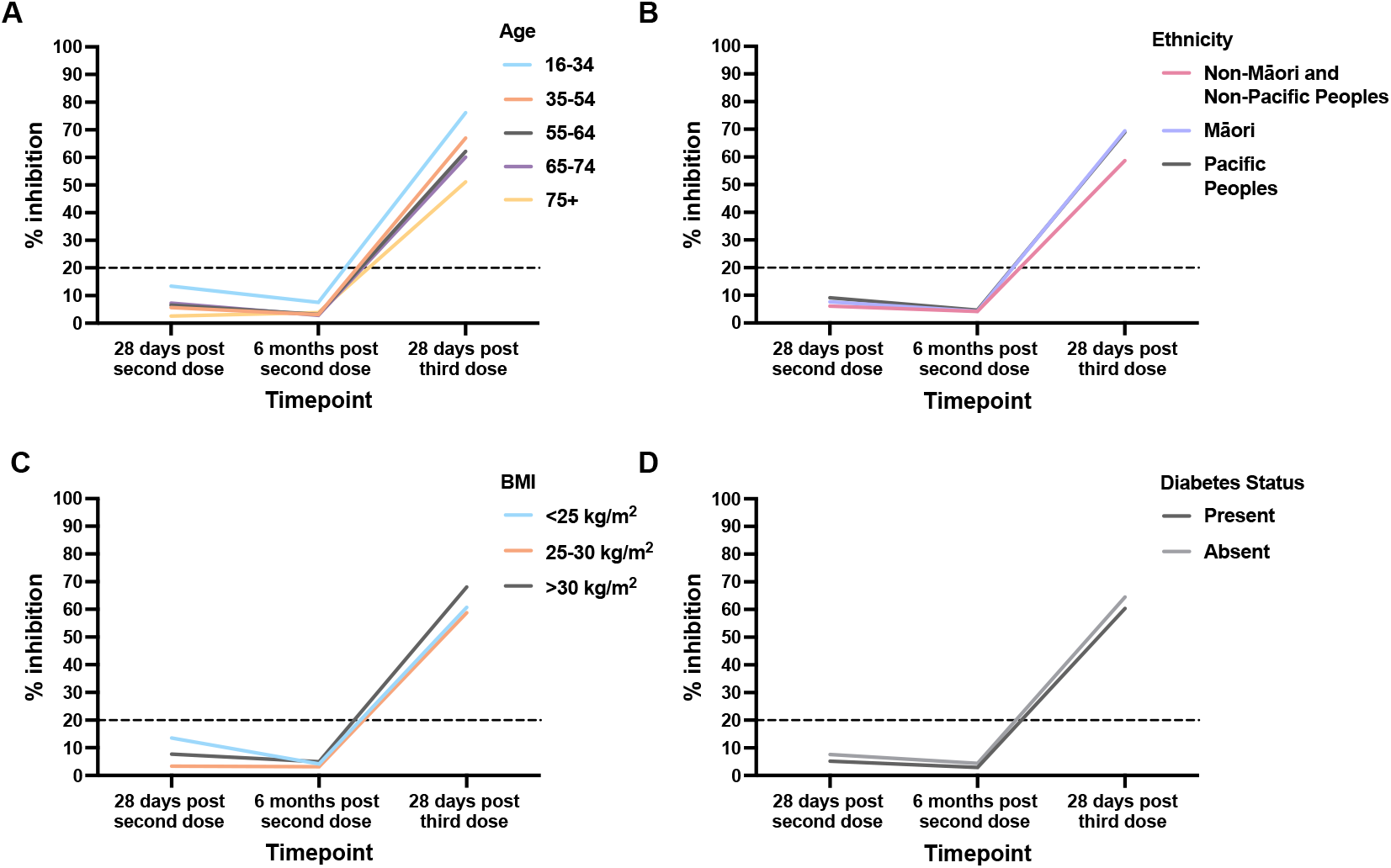
Unadjusted mean SARS-CoV-2 neutralising antibody responses to Omicron BA.1 variant, for key demographic groups, after two and three doses of BNT162b2 vaccine. Cut-off for detectable result is 20% inhibition (horizontal dotted line).

**Table 3.**
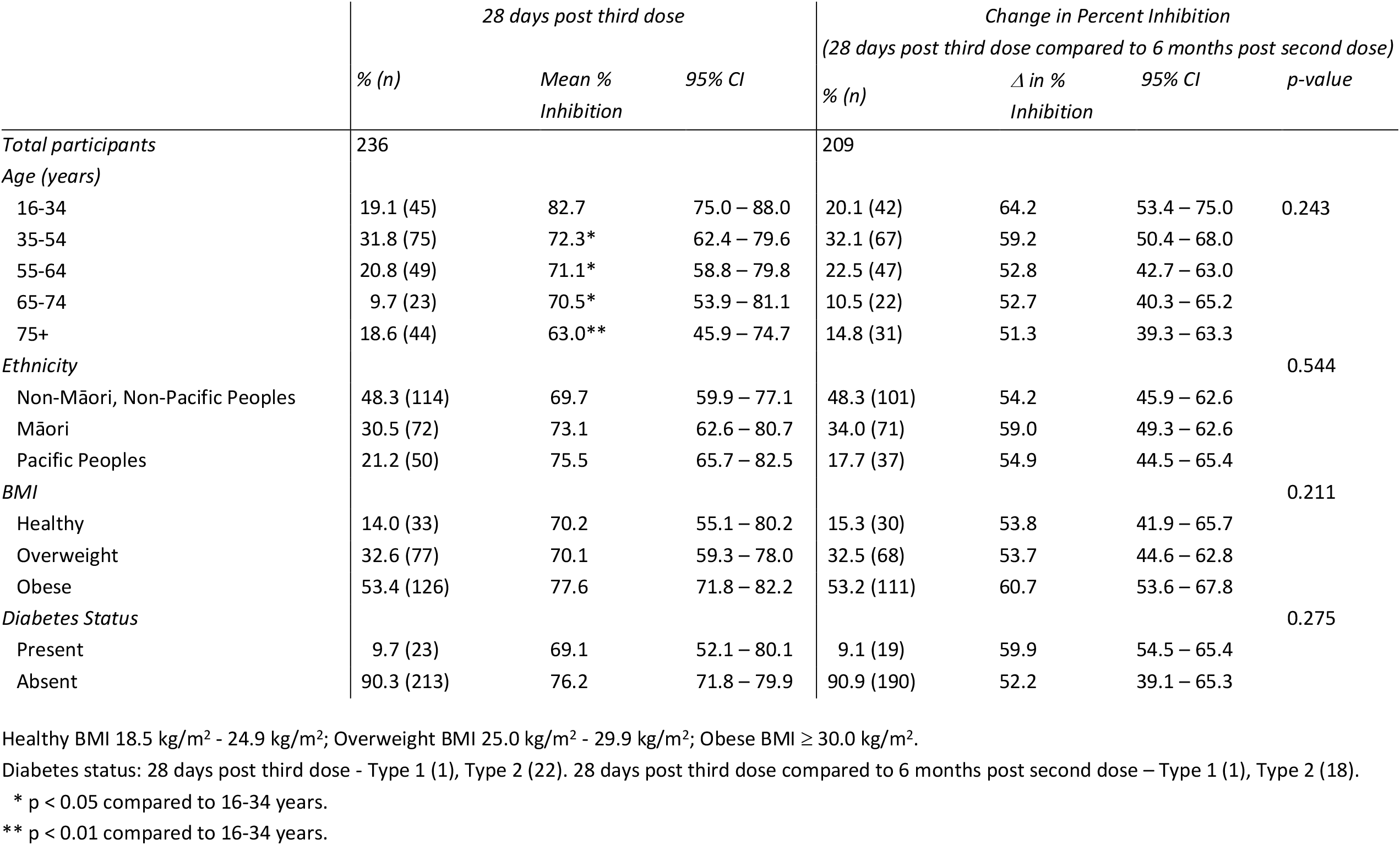
SARS-CoV-2 Omicron BA.1 neutralising antibody responses after three doses of BNT162b2 vaccine, adjusted for age, ethnicity, BMI, and diabetes status.

## Discussion

It is becoming increasingly difficult to predict population level SARS-CoV-2 immunity given variations in vaccination and prior infection history around the globe. This study cohort contributes to the growing body of international research surrounding the immunogenicity of three doses of the Pfizer-BioNTech BNT162b2 COVID-19 vaccine. It provides unique insight into rarely observed infection naïve populations, including ethnicities not explored elsewhere. Understanding the results of this study will help to inform mRNA vaccination schedules in future pandemics where the global population is naïve to a virus with relatively high rates of mutation.

An important finding from these analyses is that the third dose of Pfizer-BioNTech BNT162b2 vaccine was able to significantly increase the mean SARS-CoV-2 anti-S IgG titres of all demographic groups above titres found 28 days post dose two. While two doses in diabetics did not result in comparable titres to non-diabetics, the third vaccine dose removed this discrepancy, and is a significant finding. Furthermore, all demographic groups are afforded a substantial neutralising antibody response (over 50% neutralisation in the sVNT assay) to the Omicron BA.1 variant from the third vaccine dose that two doses were unable to provide. This is noteworthy as the Omicron BA.1 sequence is not included in the Ancestral based BNT162b2 vaccine and the vast majority of the study cohort were infection naive at the time of testing.

Older age groups, such as those aged 75 years or older, in this cohort appear to have a more complex response to the third vaccine dose. At each timepoint they have significantly lower SARS-CoV-2 anti-S IgG titres compared to younger age groups. However, like their younger comparators, they have significant improvement in GMTs after three doses compared to two doses, which mirrors results of another study [16]. Comparing GMT 6 months post second dose to 28 days post third dose, 75+ year-olds receive the greatest benefit from the third vaccine dose. However, the ability of 75+ age group to neutralise variant Omicron BA.1 is reduced compared to younger age groups.

Waning immunity was seen in this cohort with both anti-S IgG antibody levels and neutralising responses reducing over a 3-6-month period after dose two of BNT162b2. Interestingly, there was a clear consistency in waning of anti-S IgG antibody titres across age, ethnicity, and BMI which contrasts with other published data [17]. Prior international research has shown a greater rate of waning in older age groups (60 years and over)[17], as well as those with, as well ashigher BMI (>40.0 kg/m^2^after) after two doses of vaccine [18]. It is unclear if the differences with published literature are due to the unique ethnic groups within New Zealand, and this study population, the lack of circulating SARS-CoV-2 in New Zealand at the time of the study, or from differences in study design and testing.

Data on immune responses to COVID-19 vaccines in diabetics has been contradictory [19]. In contrast to other demographic subgroups in this cohort, participants with diabetes exhibited a greater waning at 6 months post second dose. Despite lower antibody levels after two doses in diabetic participants, the difference in antibody titres were corrected following a third dose, to equivalent levels as the non-diabetics in our study. This is a significant study finding and proves a third vaccine dose is of particular importance for diabetics in New Zealand.

Despite the early success and elimination of SARS-CoV-2 in New Zealand, there was a significant increase in community transmission as third doses were being administered in New Zealand, initially with the Delta variant (predominantly in the Auckland region) and then with the Omicron variant (nationwide) [3]. However, only a small proportion of our study participants had evidence of infection, therefore it was not possible to assess the impact of hybrid immunity on immunogenicity in this cohort. Nucleocapsid seroprevalence was used as a surrogate marker of previous SARS-CoV-2 infection. There are limitations in relying upon this marker as some individuals may not mount a nucleocapsid antibody response following SARS-CoV-2 infection, particularly if previously vaccinated, and these antibodies have been observed to wane over time [13, 20-22]. However, waning of anti-nucleocapsid antibodies is assay dependent, with the Roche total anti-nucleocapsid antibody assay, used in this study, reported to show high sensitivity out to at least 11.5 months post infection [23]. Participants were also screened at study commencement for nucleocapsid seroprevalence to ensure our population was infection naïve prior to participation. Therefore, the number of participants with detectable anti-nucleocapsid antibodies in this study is likely to represent the true number of study participants with prior infection, increasing validity of our results.

This study presents a unique and important perspective on response to the original BNT162b2 COVID-19 vaccine, in an infection naïve setting compared to populations with high rates of prior or intercurrent infection internationally [17, 24]. Conflicting studies indicate that the order of when infection and vaccination occur has an impact on antibody titres from vaccination, specifically infection prior to vaccination can decrease [24] or increase titres [25] and therefore protection. It is also known that natural infection with one variant, such as Omicron BA.1, may not be sufficient to protect from other variants, such as Omicron BA.2 [26]. Data from the New Zealand Ministry of Health shows that New Zealanders who had received three or more doses of BNT162b2 had a 66% reduction in the risk of death [9]. The emergent cross neutralisation from three BNT162b2 doses, as observed in this study, likely protects against the severe consequences of subsequent infections, particularly with Omicron variants. One theory for how this Omicron BA.1 neutralisation arose from a third dose, is the preferential maturation of antibodies specific to the RBD portion of the spike protein, where there are less dissimilarities between the Ancestral and Omicron variants [27].

The induction of Omicron BA.1 cross neutralisation in this cohort is consistent with previous literature that shows increased antibody titres and neutralisation after three doses against Omicron variants [28, 29] however, this is reduced in populations over 65 years of age [28]. Older groups in this study (65-74 and 75+ years-old) showed reduced antibody titres and neutralisation capacity following three doses, raising concerns that these populations are less protected from new variants. Bivalent vaccines, which encode both Ancestral and Omicron spike protein, may be able to abrogate these effects in the elderly.

Our data shows the strong ability of a third BNT162b2 vaccine dose, in a setting with limited natural infection and hybrid immunity, to produce a neutralising response against the Omicron BA.1 variant regardless of age, BMI, ethnicity, or presence or absence of diabetes. Ka Mātau, Ka Ora is an ongoing cohort study and will continue to monitor the differences seen in immunogenicity, as participants develop hybrid immunity following infection with different variants of concern, and antibody waning alongside cellular immune responses.

## Data Availability

All data produced in the present study are available upon reasonable request to the authors.

## Acknowledgements

We would like to acknowledge the study participants and the study teams at both sites for their commitment to the study, including Michelle Sampson (Lead Clinical Coordinator Lakeland Clinical Trials), Nina Linton (Study Coordinator Lakeland Clinical Trials), Gary Lees (Director of Nursing and Midwifery Lakes District Health Board), Ali Maoate (Study Coordinator Southern Clinical Trials), Dr Monica Nua George (Medical Director Etu Pasifika). Dr Api Talemaitoga provided input on study design and engaging Pacific peoples in the study. The Te Urungi Māori Advisory group of Malaghan Institute of Medical Research (MIMR) provided input on engaging Māori in the study. Mr John Treanor provided Māori consultation for the Lakeland Clinical Trials site. Dr Matea Gillies was the Māori advisor, Ngai Tahu. Dr Peter McIntyre provided peer review of the study protocol for HDEC review. Giulia Giunti and Bethany Andrews at MIMR provided operational support for the study. Ciara Ramiah provided laboratory assistance at the University of Auckland.

## Author Contributions

MB, FP, JU, KW, and GLG developed the study concept. FP, MB, and BL developed the study protocol. Study conduct, data and sample collection was overseen by MW and SC. BL, CH, and KG oversaw study coordination and data management. NM, RM, and AP ran and analysed the surrogate viral neutralisation assay. CF, CH, and BL lead the data and statistical analysis. BL, CH, CF, and MB wrote the report. Each author provided valuable feedback and revisions to the publication.

## References

1. World Health Organisation. WHO Director-General’s statement on IHR Emergency Committee on Novel Coronavirus (2019-nCoV). 2020 [cited 2022 25 November]; Available from: https://www.who.int/director-general/speeches/detail/who-director-general-s-statement-on-ihr-emergency-committee-on-novel-coronavirus-(2019-ncov).

2. New Zealand Government. Prime Minister: COVID-19 Alert Level increased. 2020 [cited 2022 14 December]; Available from: https://www.beehive.govt.nz/speech/prime-minister-covid-19-alert-level-increased.

3. Ministry of Health, More than 40,000 booster doses given yesterday; 24 community cases; 8 in hospital. 2022, Ministry of Health: https://www.health.govt.nz.

4. Ministry of Health. COVID-19 update, 17 August. 2021 [cited 2022 14 December]; Available from: https://www.health.govt.nz/news-media/news-items/covid-19-update-17-august.

5. Ministry of Health, Boosters key to protecting New Zealanders from Omicron in 2022. 2022: https://www.health.govt.nz.

6. ESR, COVID-19 Genomics Insights Dashboard (CGID) #14 - 7 July 2022. 2022, ESR

7. ESR, COVID-19 Genomics Insights Dashboard (CGID) #36 - 17 March 2023. 2023, ESR.

8. Harrison, S.L., et al., Comorbidities associated with mortality in 31,461 adults with COVID-19 in the United States: A federated electronic medical record analysis. PLOS Medicine, 2020. 17(9): p. e1003321.

9. Public Health Agency, COVID-19 Mortality in Aotearoa New Zealand: Inequities in Risk. 2022, Ministry of Health: Wellington: Ministry of Health.

10. Priddy, F.H., et al., Immunogenicity of BNT162b2 COVID-19 vaccine in New Zealand adults. Vaccine, 2022. 40(34): p. 5050–5059.

11. Ministry of Health, HISO 10001:2017 Ethnicity Data Protocols. 2017: ellington: Ministry of Health.

12. Ministry of Health. Body Size. 2018 [cited 2022 25 November]; Available from: https://www.health.govt.nz/our-work/populations/maori-health/tatau-kahukura-maori-health-statistics/nga-tauwehe-tupono-me-te-marumaru-risk-and-protective-factors/body-size.

13. Priddy, F.H., et al., Immunogenicity of BNT162b2 COVID-19 vaccine in New Zealand adults. 2022(1873–2518 (Electronic)).

14. Meyer, B., et al., Validation and clinical evaluation of a SARS-CoV-2 surrogate virus neutralisation test (sVNT). Emerg Microbes Infect, 2020. 9(1): p. 2394–2403.

15. Statistics New Zealand. A case study of 2018 Census ethnic group summaries: MELAA. 2018 [cited 2022 14 December]; Available from: https://www.stats.govt.nz/news/ethnic-group-summaries-reveal-new-zealands-multicultural-make-up/.

16. Falsey, A.R., et al., SARS-CoV-2 Neutralization with BNT162b2 Vaccine Dose 3. New England Journal of Medicine, 2021. 385(17): p. 1627–1629.

17. Pérez-Alós, L., et al., Modeling of waning immunity after SARS-CoV-2 vaccination and influencing factors. Nature Communications, 2022. 13(1): p. 1614.

18. van der Klaauw, A.A., et al., Accelerated waning of the humoral response to SARS-CoV-2 vaccines in obesity. medRxiv, 2022: p. 2022.06.09.22276196.

19. Boroumand, A.B., et al., Immunogenicity of COVID-19 vaccines in patients with diabetes mellitus: A systematic review. Front Immunol, 2022. 13: p. 940357.

20. Whitcombe, A.L., et al., Comprehensive analysis of SARS-CoV-2 antibody dynamics in New Zealand. Clin Transl Immunology, 2021. 10(3): p. e1261.

21. McGregor, R., et al., The persistence of neutralising antibodies up to 11 months after SARS CoV-2 infection in the southern region of New Zealand. N Z Med J, 2022. 135(1550): p. 162–166.

22. Craigie, A., et al., SARS-CoV-2 antibodies in the Southern Region of New Zealand, 2020. Pathology, 2021. 53(5): p. 645–651.

23. Coppola, A., et al., Durability of Humoral Immune Responses to SARS-CoV-2 in Citizens of Ariano Irpino (Campania, Italy): A Longitudinal Observational Study With an 11.5-Month Follow-Up. Front Public Health, 2021. 9: p. 801609.

24. Anderson, M., et al., SARS-CoV-2 Antibody Responses in Infection-Naive or Previously Infected Individuals After 1 and 2 Doses of the BNT162b2 Vaccine. JAMA Network Open, 2021. 4(8): p. e2119741–e2119741.

25. Carazo, S., et al., Estimated Protection of Prior SARS-CoV-2 Infection Against Reinfection With the Omicron Variant Among Messenger RNA–Vaccinated and Nonvaccinated Individuals in Quebec, Canada. JAMA Network Open, 2022. 5(10): p. e2236670–e2236670.

26. Trombetta, C.M., et al., Immune response to SARS-CoV-2 Omicron variant in patients and vaccinees following homologous and heterologous vaccinations. Communications Biology, 2022. 5(1): p. 903.

27. Cameroni, E., et al., Broadly neutralizing antibodies overcome SARS-CoV-2 Omicron antigenic shift. Nature, 2022. 602(7898): p. 664–670.

28. Lassaunière, R., et al., Neutralizing Antibodies Against the SARS-CoV-2 Omicron Variant (BA.1) 1 to 18 Weeks After the Second and Third Doses of the BNT162b2 mRNA Vaccine. JAMA Network Open, 2022. 5(5): p. e2212073–e2212073.

29. Haveri, A., et al., Neutralizing antibodies to SARS-CoV-2 Omicron variant after third mRNA vaccination in health care workers and elderly subjects. European Journal of Immunology, 2022. 52(5): p. 816–824.

