## Supplemental Files for "Robust immunogenicity of a third BNT162b2 vaccination against SARS-CoV-2 Omicron variant in a naïve New Zealand cohort"

**Supplementary Table 1**

Comorbidities present in study participants separated by ethnicity, gender, and age groups.

|  |  | Ethnicity | | | Gender | | | Age (years) | | | | |
| --- | --- | --- | --- | --- | --- | --- | --- | --- | --- | --- | --- | --- |
|  | *% (n)* | *Non-Māori, Non-Pacific Peoples* | *Māori* | *Pacific Peoples* | *Female* | *Male* | *Non-Binary* | *16-34* | *35-54* | *55-64* | *65-74* | *75+* |
| Total Participants | 288 | 121 | 86 | 81 | 156 | 131 | 1 | 72 | 88 | 55 | 26 | 47 |
| Cardiac |  |  |  |  |  |  |  |  |  |  |  |  |
| Heart Failure | 1.7 (5) | 2.5 (3) | 2.32 (2) | 0.0 (0) | 1.9 (3) | 1.5 (2) | 0.0 (0) | 0.0 (0) | 0.0 (0) | 3.6 (2) | 0.0 (0) | 6.4 (3) |
| Coronary Heart Disease | 3.5 (10) | 5.0 (6) | 3.5 (3) | 1.2 (1) | 2.6 (4) | 4.6 (6) | 0.0 (0) | 0.0 (0) | 0.0 (0) | 1.8 (1) | 11.5 (3) | 12.8 (6) |
| Cardiomyopathy | 0.3 (1) | 0.0 (0) | 1.2 (1) | 0.0 (0) | 0.6 (1) | 0.0 (0) | 0.0 (0) | 0.0 (0) | 0.0 (0) | 0.0 (0) | 0.0 (0) | 2.1 (1) |
| Hypertension | 18.4 (53) | 24.0 (29) | 14.0 (12) | 14.8 (12) | 19.2 (30) | 17.6 (23) | 0.0 (0) | 2.8 (2) | 11.4 (10) | 16.4 (9) | 42.3 (11) | 44.6 (21) |
| Pulmonary |  |  |  |  |  |  |  |  |  |  |  |  |
| COPD | 1.0 (3) | 2.5 (3) | 0.0 (0) | 0.0 (0) | 1.3 (2) | 0.8 (1) | 0.0 (0) | 0.0 (0) | 0.0 (0) | 0.0 (0) | 3.8 (1) | 4.3 (2) |
| Moderate-Severe Asthma | 9.0 (26) | 46.2 (12) | 14.0 (12) | 2.5 (2) | 14.7 (23) | 2.3 (3) | 0.0 (0) | 5.6 (4) | 11.4 (10) | 9.1 (5) | 11.5 (3) | 8.51 (4) |
| Smoking/Vaping | 6.6 (19) | 9.9 (1) | 14.0 (12) | 7.4 (6) | 6.4 (10) | 6.9 (9) | 0.0 (0) | 6.9 (5) | 11.4 (10) | 5.5 (3) | 3.8 (1) | 0.0 (0) |
| Hematology/Oncology |  |  |  |  |  |  |  |  |  |  |  |  |
| Cancer | 4.9 (14) | 5.0 (6) | 8.1 (7) | 1.2 (1) | 5.1 (8) | 4.6 (6) | 0.0 (0) | 0.0 (0) | 2.3 (2) | 7.3 (4) | 11.5 (3) | 10.6 (5) |
| Endocrine |  |  |  |  |  |  |  |  |  |  |  |  |
| Diabetes | 9.7 (28) | 5.8 (7) | 4.7 (4) | 21.0 (17) | 9.6 (15) | 9.9 (13) | 0.0 (0) | 1.4 (1) | 12.5 (11) | 9.1 (5) | 19.2 (5) | 12.8 (6) |
| Overweight | 28.1 (81) | 33.9 (41) | 30.2 (26) | 17.3 (14) | 28.8 (45) | 27.5 (36) | 0.0 (0) | 20.8 (15) | 30.7 (27) | 34.5 (19) | 23.1 (6) | 29.8 (14) |
| Obesity | 55.9 (161) | 40.5 (49) | 60.5 (52) | 74.0 (60) | 55.1 (86) | 56.5 (74) | 100.0 (1) | 50.0 (36) | 64.8 (57) | 54.5 (30) | 53.8 (14) | 51.1 (24) |
| Chronic Kidney Disease | 1.0 (3) | 0.0 (0) | 1.2 (1) | 2.5 (2) | 0.6 (1) | 1.5 (2) | 0.0 (0) | 0.0 (0) | 0.0 (0) | 1.8 (1) | 3.8 (1) | 2.1 (1) |
| Liver Disease | 0.3 (1) | 9.9 (1) | 0.0 (0) | 0.0 (0) | 0.0 (0) | 0.8 (1) | 0.0 (0) | 0.0 (0) | 0.0 (0) | 1.8 (1) | 0.0 (0) | 0.0 (0) |
| ≥ 2 Comorbidities | 35.4 (102) | 36.4 (44) | 37.2 (32) | 32.1 (26) | 39.1 (61) | 31.3 (41) | 0.0 (0) | 11.1 (8) | 36.4 (32) | 40.0 (22) | 65.4 (17) | 48.9 (23) |

COPD = Chronic Obstructive Pulmonary Disease.

Comorbidities with no prevalence in our cohort: Interstitial lung disease, pulmonary hypertension, cystic fibrosis, dementia, cerebrovascular disease, sickle cell anaemia or thalassemia, solid organ or haematopoietic stem cell transplant, HIV, immunocompromised, substance abuse disorder, Down syndrome.

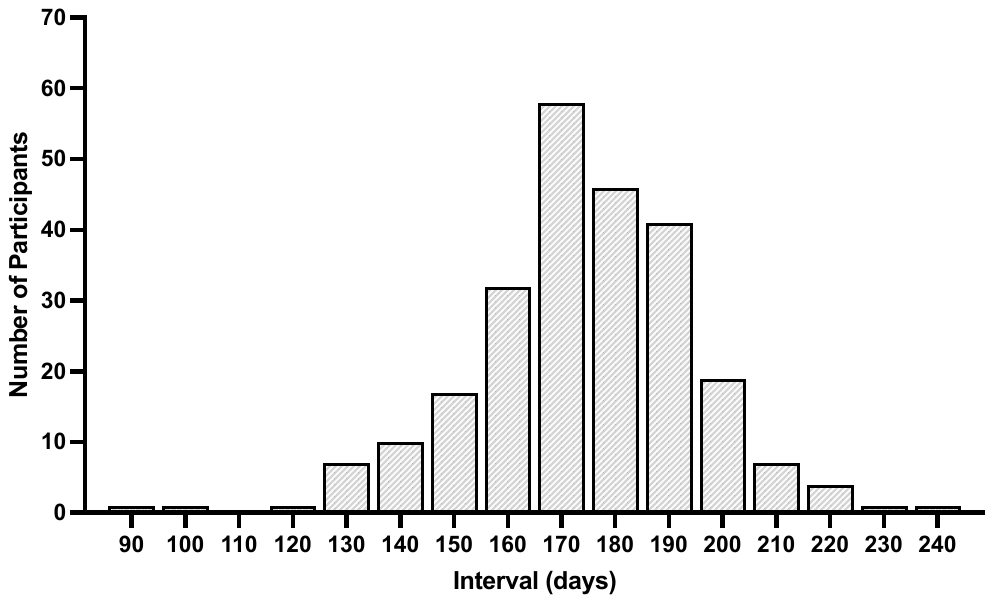

**Supplementary Figure 1.** Time interval between second and third dose of BNT162b2 vaccine.

A

A

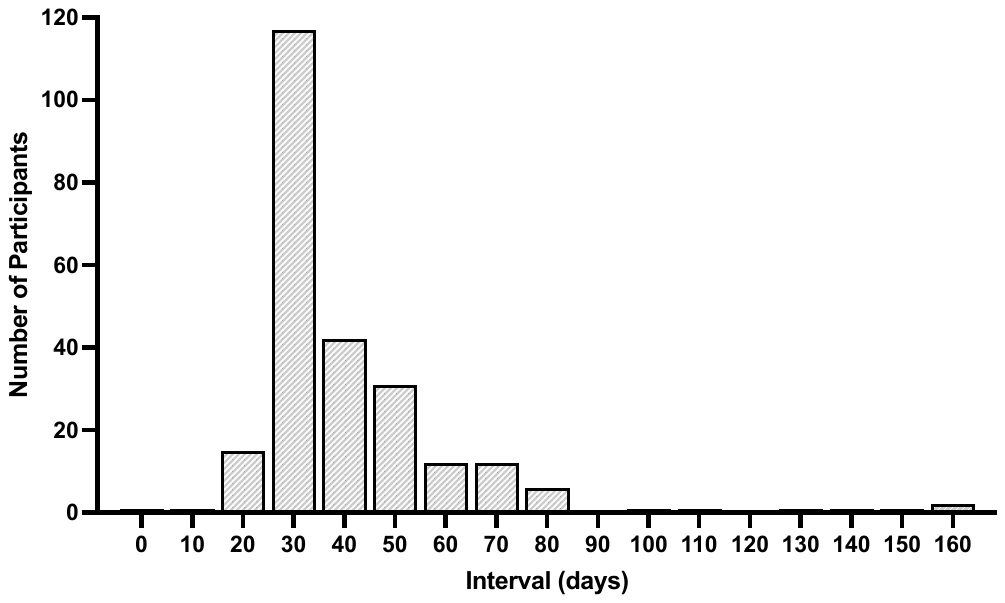

**Supplementary Figure 2.** Time interval between third dose of BNT162b2 vaccine and post vaccine sample collection for laboratory analyses.

**Supplementary Table 2**

Effect of vaccination and sampling windows as quartiles on SARS-CoV-2 neutralising, and anti-S IgG binding antibody results.

|  | *Quartiles*  *(Days)* | *6 months post second dose* | | | | *28 days post third dose* | |
| --- | --- | --- | --- | --- | --- | --- | --- |
|  |  | *Anti-S IgG*  *(AU/mL)* | *Ancestral % Inhibition* | *Beta %*  *Inhibition* | *Delta % Inhibition* | *Anti-S IgG (AU/mL)* | *Omicron % Inhibition* |
| *Interval between*  *second and*  *third vaccination* | ≤ 162 | p = 0.110 | p = 0.101 | p = 0.183 | p = 0.146 | p = 0.801 | p = 0.709 |
|  | 163 - 174 |  |  |  |  |  |  |
|  | 175 - 188 |  |  |  |  |  |  |
|  | ≥ 189 |  |  |  |  |  |  |
| *Interval between third vaccination and sample collection* | ≤ 28 | p = 0.214 | p = 0.332 | p = 0.539 | p = 0.286 | p = 0.141 | p = 0.327 |
|  | 29 - 32 |  |  |  |  |  |  |
|  | 33 - 47 |  |  |  |  |  |  |
|  | ≥ 48 |  |  |  |  |  |  |

**Supplementary Table 3**

Comparing SARS-CoV-2 Omicron BA.1 neutralising antibody responders to non-responders after two and three doses of BNT162b2 vaccine, adjusted for age, ethnicity, BMI, and diabetes status.

|  | *28 days post second dose* | | | *6 months post second dose* | | | *28 days post third dose* | | |
| --- | --- | --- | --- | --- | --- | --- | --- | --- | --- |
|  | *p-value* | *% (n) non-responders* | *% (n) responders* | *p-value* | *% (n) non-responders* | *% (n) responders* | *p-value* | *% (n) non-responders* | *% (n) responders* |
| *Total* |  | 87.1 (250) | 12.9 (37) |  | 95.8 (226) | 4.2 (10) |  | 11.9 (28) | 88.1 (208) |
| *Age (years)* | 0.010 |  |  | 0.012 |  |  | 0.004 |  |  |
| 16-34 |  | 76.1 (54) | 23.9 (17) |  | 87.7 (50) | 12.3 (7) |  | 6.7 (3) | 93.3 (42) |
| 35-54 |  | 89.8 (79) | 10.2 (9) |  | 97.2 (70) | 2.8 (2) |  | 6.7 (5) | 93.3 (70) |
| 55-64 |  | 87.3 (48) | 12.7 (7) |  | 100.0 (52) | 0.0 (0) |  | 18.4 (9) | 81.6 (40) |
| 65-74 |  | 88.5 (23) | 11.5 (3) |  | 100.0 (22) | 0.0 (0) |  | 0.0 (0) | 100.0 (23) |
| 75+ |  | 97.9 (46) | 2.1 (1) |  | 97.0 (32) | 3.0 (1) |  | 25.0 (11) | 75.0 (33) |
| *Ethnicity* | 0.291 |  |  | 0.596 |  |  | 0.503 |  |  |
| Non-Māori and Non-Pacific Peoples |  | 90.1 (109) | 9.9 (12) |  | 95.4 (103) | 4.6 (5) |  | 14.0 (16) | 86.0 (98) |
| Māori |  | 87.2 (75) | 12.8 (11) |  | 97.5 (77) | 2.5 (2) |  | 8.3 (6) | 91.7 (66) |
| Pacific Peoples |  | 82.5 (66) | 17.5 (14) |  | 93.9 (46) | 6.1 (3) |  | 12.0 (6) | 88.0 (44) |
| *BMI* | 0.215 |  |  | 0.878 |  |  | 0.500 |  |  |
| Healthy |  | 82.9 (34) | 17.1 (7) |  | 97.1 (34) | 2.9 (1) |  | 6.1 (2) | 93.9 (31) |
| Overweight |  | 92.1 (82) | 7.9 (7) |  | 96.0 (72) | 4.0 (3) |  | 11.7 (9) | 88.3 (68) |
| Obese |  | 85.4 (134) | 14.6 (23) |  | 95.2 (120) | 4.8 (6) |  | 13.5 (17) | 86.5 (109) |
| *Diabetes Status* | 0.339 |  |  | 0.312 |  |  | 0.388 |  |  |
| Present |  | 92.9 (26) | 7.1 (2) |  | 100.0 (21) | 0.0 (0) |  | 17.4 (4) | 82.6 (19) |
| Absent |  | 86.5 (224) | 13.5 (35) |  | 95.3 (205) | 4.7 (10) |  | 11.3 (24) | 88.7 (189) |

Responders were categorized as >20% inhibition against Omicron BA.1 variant in the sVNT assay.

Healthy BMI 18.5 kg/m^2^ - 24.9 kg/m^2^; Overweight BMI 25.0 kg/m^2^ – 29.9 kg/m^2^; Obese BMI ≥ 30.0 kg/m^2^.

Diabetes status: 28 days post second dose – Type 1 (2), Type 2 (26). 6 months post second dose – Type 1 (2), Type 2 (19). 28 days post third dose – Type 1 (1), Type 2 (22).
